# Investigating the impact of London’s Ultra Low Emission Zone on children’s health: Children’s Health in London and Luton (CHILL): Protocol for a prospective parallel cohort study

**DOI:** 10.1101/2021.02.04.21251049

**Authors:** Grainne Colligan, Ivelina Tsocheva, James Scales, Jasmine Chavda, Rosamund Dove, Harpal Kalsi, Helen Wood, Louise Cross, Chris Newby, Amy Hall, Mia Keating, Kristian Petrovic, Bill Day, Cheryll Crichlow, Amanda Keighley, Ann Thomson, Florian Tomini, Fran Balkwill, Borislava Mihaylova, Jonathan Grigg, Esther van Sluijs, Monica Fletcher, Mark Mon-Williams, John Wright, Sean Beevers, Gurch Randhawa, Sandra Eldridge, Aziz Sheikh, James Gauderman, Frank Kelly, Ian S Mudway, Christopher J Griffiths

## Abstract

**Introduction:** Air pollution harms health across the life course. Children are at particular risk of adverse effects during development, which may impact on health in later life. Interventions that improve air quality are therefore urgently needed not only to improve public health now, but to prevent longer-term increased vulnerability to chronic disease. Low Emission Zones are a public health policy intervention aimed at reducing traffic-derived contributions to urban air pollution, but evidence that they deliver clear health benefits is lacking. We established a natural experiment design study (CHILL: Children’s Health in London and Luton) to evaluate the impacts of the introduction of London’s Ultra Low Emission Zone (ULEZ) on children’s health.

**Methods and analysis:** CHILL is a prospective two-arm parallel longitudinal cohort study of children aged 6-9 years, attending primary schools in Central London (the focus of the first phase of the ULEZ) and Luton (a comparator site). The primary goal of the study is to examine the impact of changes in annual air pollutant exposures as oxides of nitrogen, nitrogen dioxide, particulate matter less than 2.5microns and 10microns (NOx, NO_2_, PM_2.5_, PM_10_ respectively) across the two sites on lung growth, measured as forced expiratory volume in one second (FEV_1_) and forced vital capacity (FVC), over four consecutive years. Secondary outcomes being investigated include a range of respiratory health indicators as well as inequality and health economic measures.

**Ethics and dissemination:** Ethics approval has been given by Queen Mary University of London Research Ethics Committee (ref 2018/08). Dissemination will target audiences through a variety of channels, including research papers, conference and media presentations, web summaries and social media. CHILL is funded by National Institute for Health Research (NIHR) Public Health Research (Ref 16/139/09) with additional funding by Natural Environment Research Council, NIHR CLAHRC North Thames, NIHR ARC North Thames, and the Mayor of London. ClinicalTrials.gov: NCT04695093

**Strengths and limitations of this study:** *Strengths:*

- CHILL uses a prospective parallel cohort design, allowing robust conclusions to be drawn on the impact of the ULEZ - a major city-wide air quality mitigation strategy - on air quality and children’s respiratory health.
- CHILL study cohorts include children from large and ethnically diverse populations living in urban areas characterised by poor air quality.

*Limitations:*

- Attrition of study cohort population over time, although this has been accounted for in the original design of the study.
- Potential diminution of the ULEZ air pollution signal due to pre-compliance with ULEZ restrictions in the run up to the introduction of the scheme in Central London on the 8^th^ April 2019, and minor impacts of other pollution mitigation measures.
- Added complexity of accounting for effects of COVID-19 and related lockdowns on traffic flows, air quality and children’s health.

## Introduction

Air pollution is associated with adverse health effects across the life course and substantial health inequalities.^1 2^ Children are particularly vulnerable,^1 3^ with adverse effects observed on developmental trajectories and long-term health, including poor birth outcomes,^4^ stunted lung growth,^5^ delayed cognitive development^6^ and increased incidence of psychiatric disorders.^7^ Exposure to air pollution during infancy and childhood is associated with increased risk in later life of asthma, pneumonia and chronic obstructive pulmonary disease (COPD).^8^ Even exposures below legal limit values are associated with disability, disease and death in childhood.^1^

Cohort studies have played a key role in identifying these impacts. The ESCAPE meta-analysis of five European birth cohorts showed poor air quality was associated with reduced lung function in pre-adolescent children.^9^ The California Children’s Health Study (CHS) showed clinically important deficits of lung growth and function in adolescents.^10–13^ Whilst these studies have illustrated associations between adverse responses and air pollutant exposures, it is also notable that downward trends in air pollution have also been shown to deliver health improvements. This was illustrated by the successive children’s cohorts within the CHS (between 1994-2011) where the proportion of adolescents with clinically significant deficits in lung function fell as air quality in California improved.^14^

There is therefore an urgent need to identify the most impactful policy interventions that improve air quality and deliver health benefits. Low Emission Zones (LEZ), which restrict the entry of polluting vehicles to urban areas, and related Clean Air Zones, have become the dominant public health policy intervention in the effort to improve air quality across Europe.^15 16^ To date, studies evaluating their impact have shown variable effects on road use,^16^ exhaust emissions, and air quality, with few identifying clear health benefits.^5 17–24^ A systematic review which evaluated the impact of air quality strategies across Europe on health and health inequalities found negligible effects on respiratory symptoms^23^ with only one of 15 studies identified gathering health data directly from individuals.^24^ There was limited evidence for impact of LEZs on air quality in five EU countries (Denmark, the Netherlands, Germany, Italy and the UK) though the health impact of these changes was not addressed.^25^

London implemented its first LEZ from 2008 to 2012, with phased tightening of emission standards in 2008 and 2012. We evaluated its impact, using a sequential cross-sectional design, on the health of 2,297 east London primary school children,^5 26^ finding small improvements in air quality (most clearly NO_2_ reductions at the roadside), but no convincing health benefit: such as improvements in lung function or respiratory symptoms. However, over the study period, we did identify significant deficits in participating children’s lung capacity of between 5-10%, associated with exposures to traffic-related pollutants.

In 2019, London began implementing a second LEZ with more ambitious air quality targets, termed the ‘Ultra Low Emission Zone’ (ULEZ).^27^ Funded by NIHR Public Health Research, we established the Children’s Health in London and Luton (CHILL) study, a natural experiment to determine whether the impact of the ULEZ on improving air quality has a corresponding impact on children’s respiratory health (NIHR PHR 16/139/01). CHILL is a prospective two-arm parallel longitudinal cohort study of children aged 6-9 years, attending primary schools in the central area of London (the focus of the ULEZ first phase) and Luton (a comparator site). The primary goal of the study is to determine the impact of annual air pollutant exposures on lung growth (measured as FEV_1_ and FVC, post bronchodilator), over four consecutive years. These objectives were further enhanced prior to the initiation of the study to consider the impacts of the ULEZ implementation on physical activity, travel behaviours and obesity using funding from the NIHR funded Collaboration for Leadership in Applied Health Research and Care (CLAHRC) and the Applied Research Collaboration (ARC), North Thames.

By collecting sequential annual lung function, respiratory symptoms data and information on health care use from participants at both locations and relating these to annual and monthly modelled exposures to criterion air pollutants: NO_2_, ozone (O_3_) and particulate matter (both PM of less than 5 and 10 microns, PM_2.5_ and PM_10_ respectively) over the four years of the study, the proposed work will provide valuable insights into the effectiveness of air quality regulatory action on children’s health. CHILL meets all the criteria for a high-quality natural experiment evaluation including: prospective design, comparator site, large representative population sample, detailed air quality measurements, health record data, with the potential for downstream mechanistic assessments through additional funding initiatives. Our findings will add needed evidence on how pollution control strategies affect children’s health, and will inform future efforts by cities in the UK, Europe and globally to implement equally robust and ambitious air pollution interventions to achieve health benefits.

### Primary research question

- Does the implementation of London’s ULEZ improve lung function and pulmonary growth trajectories in children of primary school age?

### Secondary research questions

- To what extent does ULEZ implementation improve air quality, specifically reductions in the emissions from diesel vehicles?
- Does ULEZ implementation result in a reduction in respiratory and allergic symptoms, and respiratory infections in primary school aged children?
- Does implementation of the ULEZ encourage increased outdoor physical activity, alter travel behaviours and impact on children’s weight and risk of obesity?
- Does the ULEZ deliver measurable benefits in the perception of quality of life?
- Does the ULEZ reduce health care use and associated costs?

## Methods and analysis

### Study design

The CHILL study is a prospective parallel longitudinal cohort study performed over 4 years at two separate locations, one impacted by the ULEZ road traffic management scheme (London) and one not (Luton). The full study design is outlined in Figure 1. CHILL opened to recruitment in June 2018 to capture a year of pre-ULEZ baseline data.

**Figure 1:**
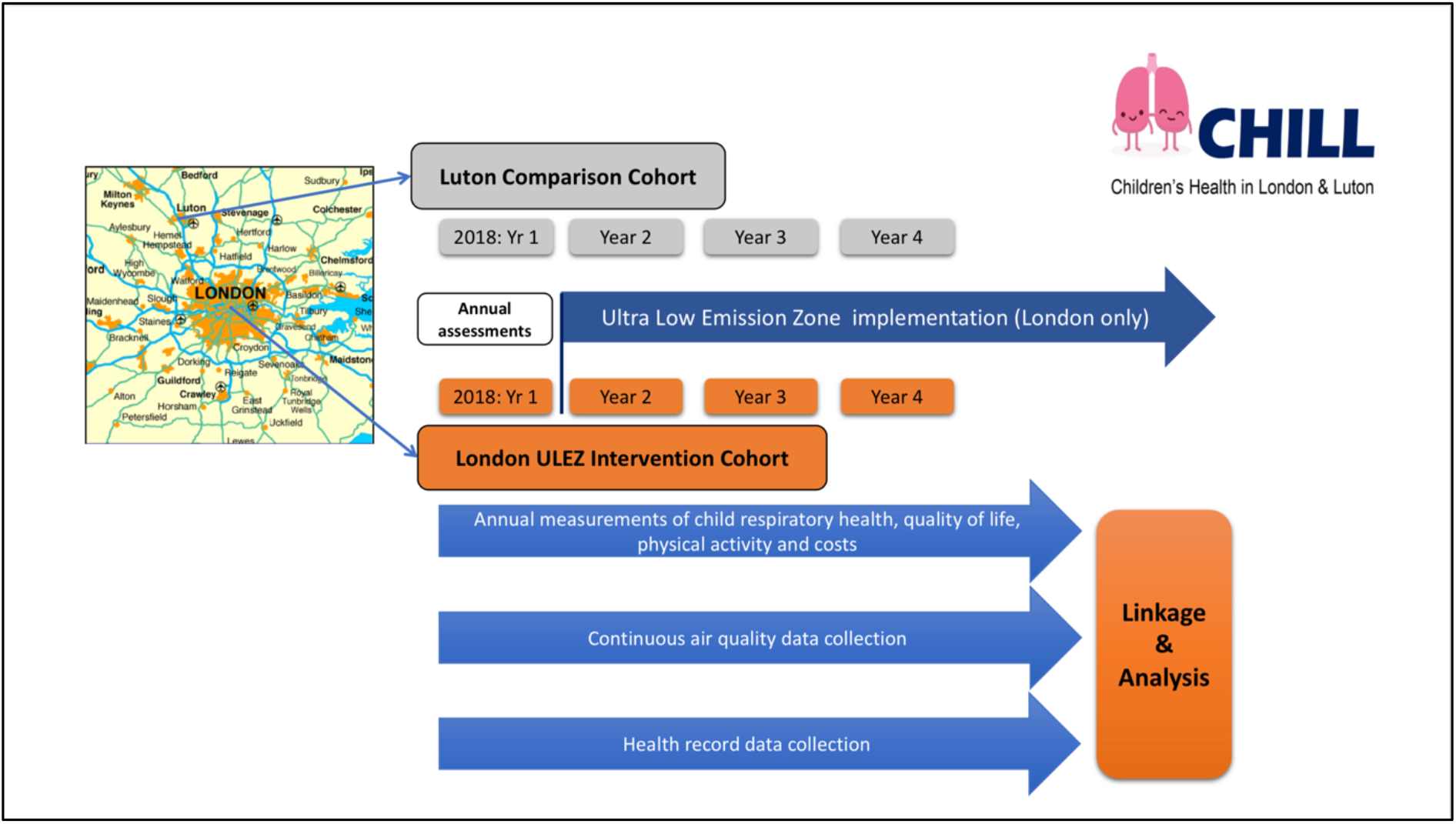
Study scheme diagram.

### Study Population

Children (age 6-9 years) were recruited from 40 London primary schools (years 2, 3, 4) with a catchment area within and bordering the Central London ULEZ area, and 40 primary schools (year 2, 3, 4) from the Boroughs of Luton and Dunstable. The Luton area was chosen as a suitable comparison site to Central London due to its broadly similar air quality, demography, and levels of socio-economic deprivation. Whilst it is subject to projected air quality improvements through national government policy,^15^ it does not have the ambitious local scale policies being enacted within London. It is also sufficiently distant from London to be free from risk of contamination by effects of the ULEZ. School and participant eligibility criteria are summarised in Table 1.

**Table 1.** Eligibility Criteria.

|  | **Primary schools** | **Individuals** |
| --- | --- | --- |
| **Inclusion criteria** | Any school located within, or whose catchment area includes the Central London ULEZ; or within the Boroughs of Luton and Dunstable. | All children attending a study school, in years 2, 3, or 4 at study inception and not meeting exclusion criteria. |
| **Exclusion criteria** | Primary schools that are not within the boundaries outlined above. | Children with learning or physical disabilities sufficient for them to be unable to give informed assent to the study, or to carry out study procedures.<br>Children with major lung disease (not including asthma). |

### Outcome measures

- **Primary**: Lung function growth (post-bronchodilator forced expiratory volume in one second, FEV_1_, per year)
- **Secondary**: Air quality, forced vital capacity (FVC) growth per year, respiratory symptoms, respiratory infections, physical activity, quality of life, health care use, costs.

### Study Interventions

#### London Ultra Low Emission Zone

The Central London ULEZ, implemented on April 8^th^ 2019 uses number plate recognition technology and a daily penalty charge notice issued for vehicles entering this central zone not meeting the set standards – as outlined below in Figure 2. The ULEZ will subsequently be extended to the North Circular Road (A406) and South Circular Road (A205) in March 2021 (orange zone Figure 2), with a London-wide ULEZ for heavy duty vehicles (HDV) within the current LEZ boundary (green zone Figure 2).

**Figure 2:**
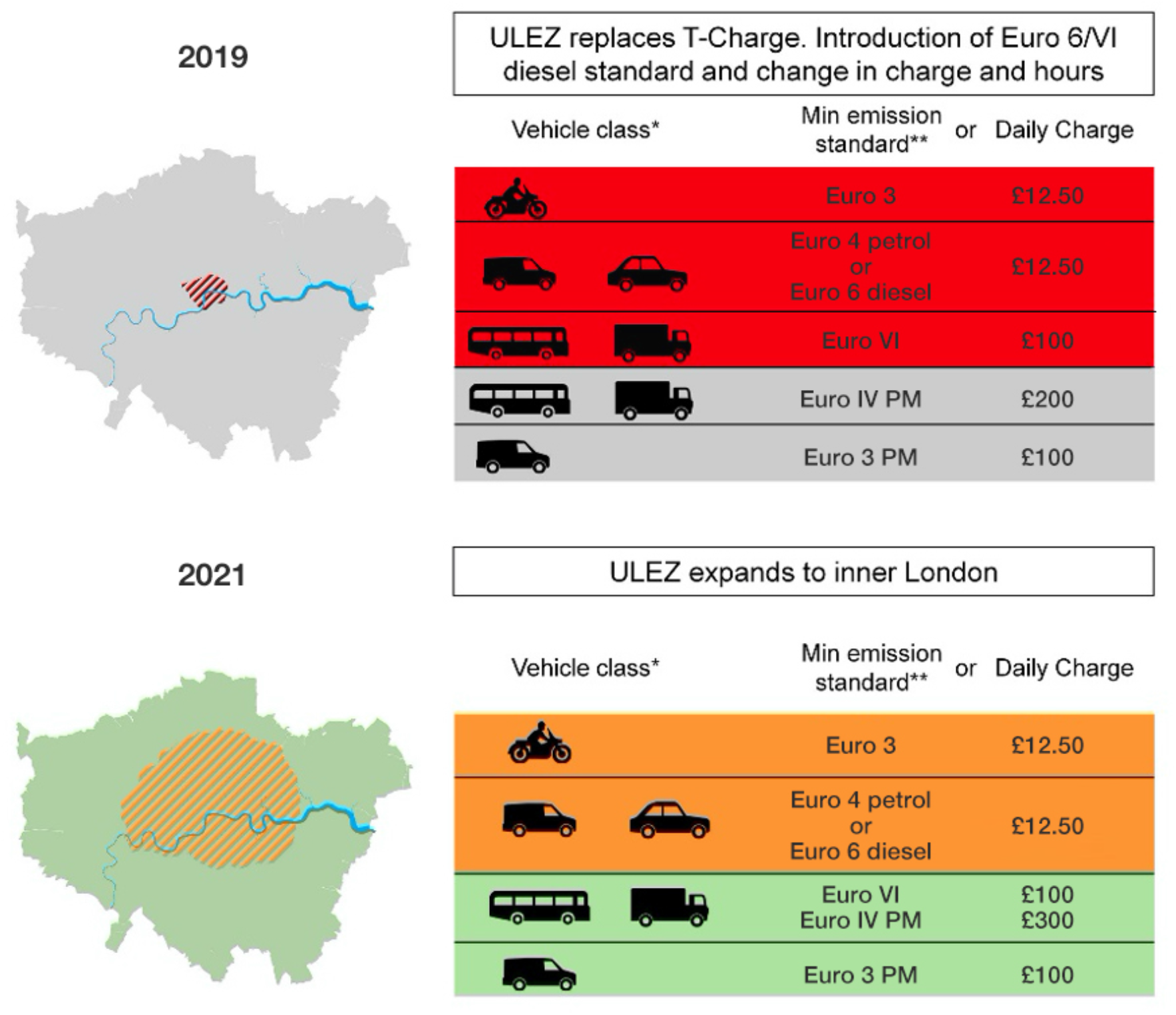
ULEZ configuration during phase 1 (2019) and the subsequent extension, scheduled for 2021. *Vehicle class is indicative only; **minimum emission standard is for NOx and PM unless otherwise stated.

### Luton: comparison site

Luton’s air quality is influenced by several factors: the presence of major industry (including a motor industry), the transecting M1 motorway and A505, and a rapidly expanding international airport, all bringing significant traffic flows into and through the town. Luton has no plans for a Low Emission or Clean Air Zone. It has three designated Air Quality Management Areas. Planned interventions for air quality improvement include a busway, car sharing, public information and advice systems, and provision of charging points for electric vehicles.

### Sample size

We designed our study to achieve 90% power at a 0.05 significance level for detecting a 15 ml difference in FEV_1_ growth per year between the Central ULEZ and Luton comparison cohorts. This design requires a total of 3,200 children, comprising 1,600 in the London cohort and 1,600 in the Luton cohort, with allowance for expected attrition as described below. To achieve this goal, we are recruiting 40 schools/arm with 40 children from any of school years 2, 3, or 4. Power calculation was carried out through simulation and testing on the interaction of FEV1 and time over the 4 measurements to obtain a lung growth (FEV_1_ per year) difference between London and Luton

### Assumptions

- Adjustment for clustering of lung function outcomes within schools, by inflating sample size using an intra-class correlation (ICC) for FEV_1_ in schools calculated from our original ULEZ study (ICC=0.001)
- 70% success rate for a valid FEV_1_ reading in children in school years 2 and 3
- 20% attrition per year of follow up, reflecting children moving schools or withdrawal
- 30% inflation to enable subgroup analyses

### Recruitment

#### School recruitment

All schools meeting the inclusion criteria were invited to take part, and made aware of the study through local media and contact with local leaders and parent groups. An invitation email with a link to a short film summarising the study,^28^ was followed up by a call from the study Chief Investigator.

#### Child recruitment and consent

Consent forms were completed by children’s caregivers at home, and require opt-in to study components, including elements of the health assessments and access to GP health records and Hospital Episode Statistics data. The recruitment and consent procedure followed a “school bag” approach, facilitated by school staff, Figure 3.

**Figure 3.**
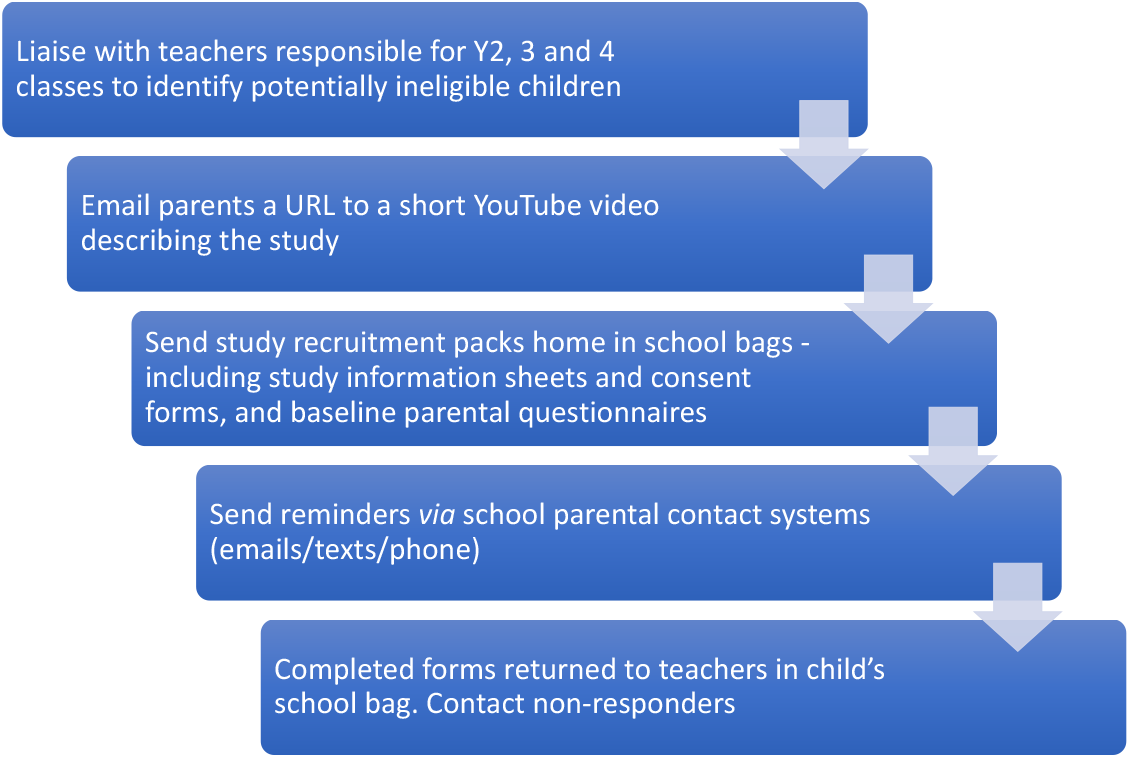
Child recruitment process.

#### Patient and Public Involvement

Patient and public involvement (PPI) is integral to the CHILL study and makes formal and informal contributions. Study design was informed by consultation with parents, headteachers, children from study areas, and community groups such as ‘Mums for Lungs’.

The CHILL Study, as a progression of previous research projects,^5^ has an established network of interested public who willingly joined the CHILL study dedicated PPI group. The group aims to ensure that the perspectives and welfare of the participant children, caregivers and schools remain at the centre of the study throughout. The PPI group: i) provides comment and advice on study materials; ii) supports recruitment and retention in the study; iii) advises on dissemination of progress and findings and iv) provides representatives who are members of the Project Management Group (PMG) and Independent Study Steering Committee (ISC).

Informal PPI between the study team and school staff, children and caregivers is ongoing throughout the lifetime of the project.

#### Science Outreach Education

Central to the CHILL study community outreach strategy is to engage children from participating schools as active participants in science and health research. Most study schools are located in areas of socioeconomic deprivation with high proportions of families from Black, Asian and ethnic minority heritages. To this end, interactive science outreach sessions addressing air pollution and health are developed and delivered by Centre of the Cell, Queen Mary University of London’s (QMUL) Science Education Centre.^29^ A learning outreach officer delivers the outreach sessions to all school classes participating in the study, regardless of how many individual children are taking part in health assessments. Children and class teachers are invited to provide feedback on the sessions to be taken into consideration for future science session planning. The intention is each year to deliver a fresh outreach session addressing a different aspect of air pollution and health (respiratory; genetic; brain and cognition) that links to the CHILL study goals.

#### Data collection methods

Using different formats and measurement tools (Table 2), data are collected in two ways: 1) directly from children at health assessments during school visits; and 2) from the questionnaire filled out by caregivers returned *via* the school bag approach.

**Table 2:** Outcomes and Data Collection.

| Type | Outcome | Method | Year 1 | Year 2 | Year 3 | Year 4 | Recording |
| --- | --- | --- | --- | --- | --- | --- | --- |
| Primary outcome | FEV<sub>1</sub> | Annual health assessment: spirometry | school visit | school visit | school visit | school visit | Spirometer download |
| Primary exposures | Air quality: NO, NO<sub>2</sub>, O<sub>3</sub>, PM<sub>10</sub>, PM<sub>2.5</sub> | Examination of long-term pollution trends over the study period in study areas | LAQN* and AURN** | LAQN and AURN | LAQN and AURN | LAQN and AURN | LAQN and AURN |
| Secondary outcome | FVC and other spirometric variables | Annual health assessment: spirometry | school visit | school visit | school visit | school visit | Spirometer download |
| Secondary outcome | Physical activity and GPS tracking | Annual health assessment: accelerometer and GPS tracker | school visit | school visit | school visit | school visit | Accelerometer and GPS download |
| Demography / potential confounding variables |  | Parent-completed questionnaire: (paper or web) | school bag prior to school visit | school bag prior to school visit | school bag prior to school visit | school bag prior to school visit | Clinical Record Form |
| Secondary outcome | School absence; work absence | Parent-completed questionnaire: (paper or web) | school bag prior to school visit | school bag prior to school visit | school bag prior to school visit | school bag prior to school visit | Clinical Record Form |
| Secondary outcome | Respiratory and allergy symptoms (ISAAC)*** | Parent-completed questionnaire: (paper or web) | school bag prior to school visit | school bag prior to school visit | school bag prior to school visit | school bag prior to school visit | Clinical Record Form |
| Secondary outcome | Quality of Life (CHU9D)**** | Parent-completed questionnaire: (paper or web) | school bag prior to school visit | school bag prior to school visit | school bag prior to school visit | school bag prior to school visit | Clinical Record Form |
| Secondary outcome | Non-NHS costs | Parent-completed questionnaire: (paper or web) | school bag prior to school visit | school bag prior to school visit | school bag prior to school visit | school bag prior to school visit | Clinical Record Form |
| Secondary outcome | Respiratory infection Health care use NHS costs | Data extraction from GP health records HES***** data linkage | Gathered after year 4 school visits complete | Gathered after year 4 school visits complete | Gathered after year 4 school visits complete | Gathered after year 4 school visits complete | Clinical Record Form |

\*LAQN: Local Air Quality Network
\*\*AURN: Automatic Urban & Rural Network
\*\*\*ISAAC: International Study of Asthma and Allergies in Childhood Questionnaire
\*\*\*\*CHU9D: Child Health Utility 9D Questionnaire
\*\*\*\*\*HES: Hospital Episode Statistics

#### Annual health assessments

Annual health assessments take place at each school as far as possible during the same calendar month of each year. The study team assess children in groups of four or five in a suitable room identified by the class-teacher, using standard protocols. Assessments take about 35 minutes per child and include:

- Height, weight, body mass index (BMI)
- Pre- and post-bronchodilator spirometry
- Provision of Actigraph^30^ physical activity accelerometer and where resources allow, a Global Position Satellite (GPS) monitor
- Annual study questionnaire for home completion by parents/guardians.

##### Spirometry

Following height (sitting and standing) and weight measurements, lung function is measured before and 15 minutes after bronchodilation, according to European Thoracic Society guidelines.^31 32^ Fully trained members of the study team use a Vitalograph 6000 Alpha Touch Spirometer,^33^ calibrated using three litre precision syringe at the start of each session. Children inhale four puffs (100mcg/puff) of salbutamol *via* a large volume spacer, administered by a study team member. A new or sterilised spacer is used for each child.

##### Physical activity

Each child is fitted with an Actigraph accelerometer and provided with instructions (verbal and paper) on its use. Children are requested to wear the monitor during waking hours for seven days after which the monitors are collected from the school. Children answer a question on their route to school and means of transport on the day of assessment. Children in some schools have the option of wearing a GPS monitor for a week to track the routes they take to and from school.

##### Annual study questionnaire for home completion

Caregivers are asked to complete questionnaires in year 1 (when consenting to the study) and in study years 2, 3 and 4. These comprise:

- Demography and residential history
- Parent-reported respiratory and allergy symptoms (ISAAC questionnaire)^34^
- Parent-reported paediatric QOL (CHU9D questionnaire)^35^
- Non-NHS costs due to child’s respiratory ill-health
- Child absence from school due to respiratory ill-health
- Smoking/exposure to second-hand smoke during pregnancy
- Travel choices
- Parental absence from work due to child’s respiratory ill-health

Upon receipt of a completed questionnaire, caregivers receive a £5 supermarket voucher.

#### NHS health records

Following approval by an NHS research ethics committee and with participants’ consent we will approach the Caldicott Guardians of participants’ general practices to request electronic downloads of coded electronic health record data from birth to current age, to capture lifetime respiratory health, mental health, COVID-19 tests and health care use. Data collection will take place during year 4 of the study.

#### Air pollution modelling

Exposure estimates will be produced every 20m^2^ in the UK using the CMAQ (The Community Multiscale Air Quality Modelling System)-urban model which couples the Advanced Dispersion roads Model (ADMS) with the Weather Research and Forecasting model (WRF) meteorological model and CMAQ regional scale models. A comprehensive description of the CMAQ-urban model has been published previously^36–38^ and will be employed in the present study to model NOx, NO_2_, O_3_, PM_2.5_, PM_10_, PM (Primary Organic Aerosol (POA), Secondary Organic Aerosol (SOA), nitrate, sulphate, black carbon), as well as exhaust and non-exhaust PM contributions continuously over the period 2018 – 2020 (CHILL). Model outputs will be provided at residential address level. Pollutant values are assigned to children on the basis of home and school postcodes, adjusted for estimated time spent at each.

#### Data protection and management

Participants are allocated a unique study identification number. Data collection, confidentiality, entry, security and storage and protection is managed according to principles of good clinical practice and in line with our Clinical Trials Unit protocols.

Data handling and record keeping are overseen by the Pragmatic Clinical Trials Unit (PCTU) based at QMUL. The Data Manager has developed appropriate data management strategies for the study and advises on their implementation. Advice is provided on the current regulatory framework regarding data protection and data management procedures in compliance with the Data Protection Act and trial regulations. All databases have integrated data validation checks and audit trails. The PCTU data management team advise on electronic data security. Electronic data is stored in secure trusted research environments. PCTU will also advise on data transfer, storage, back-up and archiving of data and ensure databases are regularly backed up and data safeguarded from accidental loss. Paper records, case record forms and consent forms and recruitment logs are held locally in line with governance procedures. Personal data will only be accessed and used by those members of the research team at QMUL and representatives of the sponsor who have been granted permission.

## Data analysis

### Statistical Methods

The effect of air pollution on post bronchodilator FEV_1_, and on secondary outcome variables of post bronchodilator FVC and physical activity, will be analysed longitudinally through a mixed effect model. The model will be rich enough to assess the pollution-outcome relationship over time at the child, school, and city levels, while incorporating appropriate random effects to account for clustering of measurements at these levels. The primary outcome of the study is lung growth over time *i*.*e*. FEV_1_ over time comparing the difference between Luton and London; this will be analysed using a mixed effect model adjusting for the levels of school and child if models are not saturated. This mixed-effect modelling framework will utilize all available data, including measurements made on study subjects that may have missed one or more visits, facilitating a longitudinal design in a highly transient population. Initial models will assume linear growth in lung function over the study period, but this and other model assumptions will be carefully examined and alternatives, such as a flexible-spline growth model,^10^ will be applied as needed. Primary and secondary outcome variables will be adjusted to account for gender, asthma diagnosis, ethnicity, deprivation,^39^ age, height, and BMI. Secondary covariates evaluated in sensitivity analysis include socioeconomic status, stress, and physical activity.

### Sensitivity analyses and missing data

Sensitivity analyses will be conducted to examine possible bias due to missing data by contrasting an “all children” data set to a “complete history” data set. Temporal individual and school exposure assessments will be contrasted to the rest of the cohort to assess cluster bias such as rebound lung growth and high non-traffic related air pollution change unrelated to the ULEZ.

Rigorous descriptive analyses of each continuous variable will be conducted to identify potential outliers or data collection errors. Plots of model residuals will also be inspected for suspected outliers. Should any be found, raw data will be checked for accuracy. If the data are found to be correct, a model will be fitted excluding outliers as a sensitivity analysis to assess impact on the primary model estimates and inferences.

### Additional analyses

Secondary analysis will assess the effect of the intervention on participants’ FEV_1_ and other outcomes, stratified by sex, ethnic group, and quintile of deprivation. Specifically, assessment will examine the following outcomes: change in air quality for each individual pollutant; lung growth; change in generic quality of life; change in respiratory symptoms; change in respiratory infection rates; change in health care use and change to small-area socioeconomic, population and mortality outcomes, z-scores and % predicted values of FEV_1_ and FVC adjusted for gender, age, height, ethnicity.

Effects of COVID-19 and related lockdowns include impacts on traffic flows, air quality and children’s health. We will carry out additional detailed air quality modelling and outcome assessments to disentangle the relative impacts of the ULEZ and COVID-19 on study outcomes.

### Assessment of cost-effectiveness

An economic evaluation will relate costs associated with introducing the ULEZ to the impact of air pollution on children’s lung growth, school attendance, parent work productivity and use/costs of healthcare services and quality of life. Analysis will use a cost-consequences approach, as recommended in National Institute for Health and Care Excellence (NICE) public health guidance.

Downloads of each child’s electronic health record will provide data on primary and secondary care use, while unit costs from national sources will be applied to all resource use to estimate individual-level costs. Impacts of school absence will be assessed in relation to meeting government attendance targets and consequent lost productivity/income due to work absence by parents, using appropriate assumptions, national wage rate data and a human capital approach to costing. Quality of life and quality-adjusted life years will be assessed by administering the parent completed proxy CHU 9D and its associated general population-based preference weights. The cost-consequences analysis will report means and standard deviations for all costs and outcomes for both cohorts with between cohort differences assessed using a mixed effects model with interaction effects of time and study site.

### Ethics, safety and dissemination

The study is approved by Queen Mary University of London research ethics committee (reference 2018/08). The Chief Investigator has the overall oversight, responsibility and a duty to ensure that safety monitoring and reporting is conducted in accordance with the sponsor’s requirements and that an annual progress report, including any safety issues, is submitted to the main research ethics committee and the sponsor. A progress report every six months is sent to the funder. The funder is advised of protocol amendments.

### Governance

An Independent Steering Committee (ISC) comprising six members and two PPI members monitors the study on behalf of the funder and sponsor. The ISC meets with the Chief Investigator and study team six monthly.

### Data monitoring and auditing

Monitoring and auditing of data is carried out by the PCTU Quality Assurance team.

### Dissemination plan and project outputs

The CHILL study will disseminate progress from the outset of the study, maximising public and professional awareness of the study and its relevance to public and child health. We expect to reach the following groups:

- Study participants, their families, schools and local study communities;
- The public - including voluntary organisations, charities, lay and pressure groups;
- Government - including Parliament, national, regional, local councils, international governments;
- Academia – including universities, NIHR, Royal Colleges, NICE, World Health Organization (WHO), and leading international research groups;
- Industry – vehicle and transport-related manufacturers.

The outputs needed to target these audiences vary (Figure 4).

**Figure 4:**
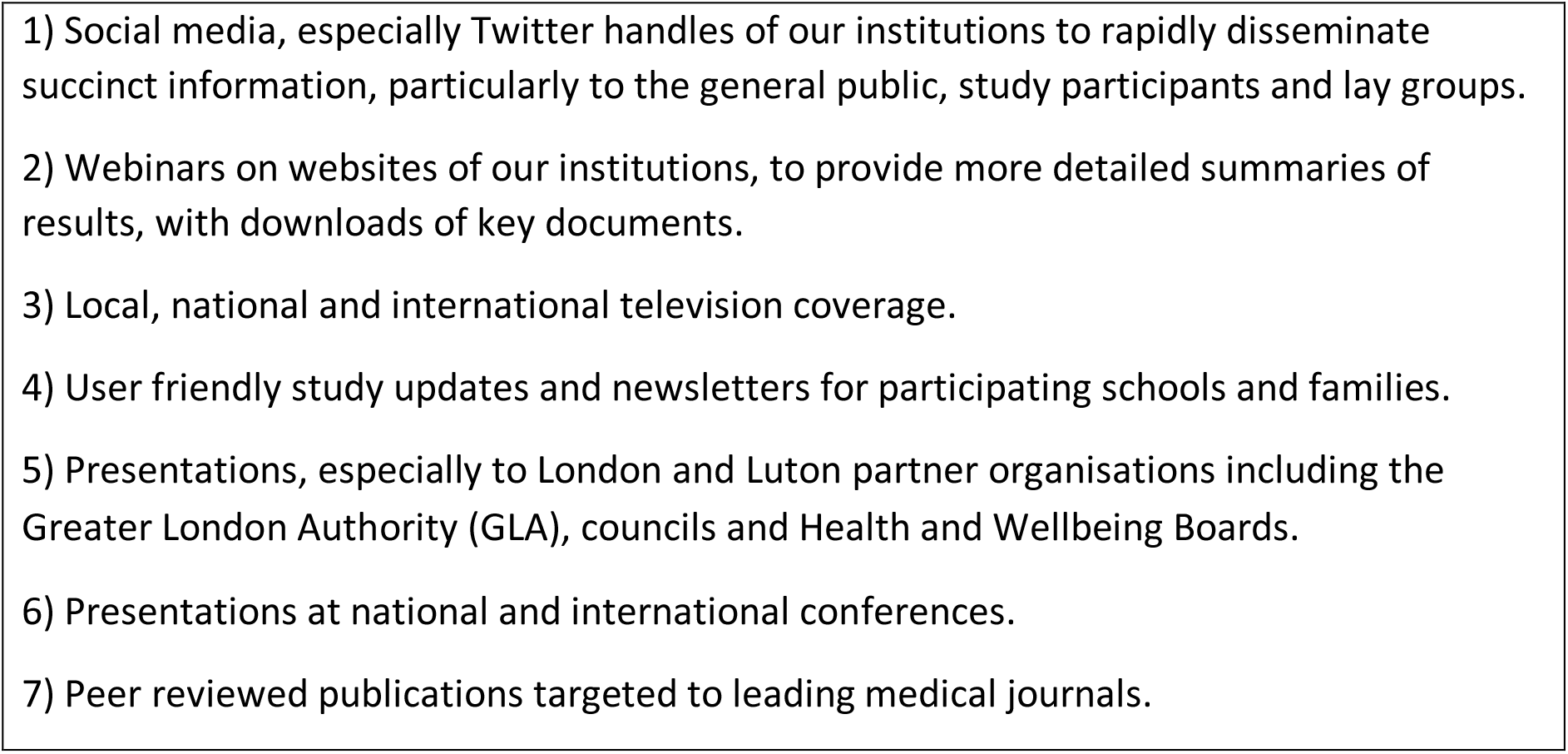
Methods of Dissemination.

## Discussion

Air pollution is regarded as the largest single environmental risk to health, with seven million deaths globally attributed annually.^40^ Environmental health and sustainable development goals identified by the WHO highlight the need for effective preventative interventions to protect children from adverse environmental exposures including air pollution.^3^ In the UK, road traffic represents the largest contributor to air pollution in urban areas,^41^ with the period between 1949 - 2012 seeing a tenfold increase in the distance travelled by the average person in their car.^2^ There is an urgent need to identify and quantify the impacts of effective air quality improvement health policy interventions. Such interventions are often costly to implement requiring large alterations in social functioning and urban infrastructure. It is essential therefore that evaluations deliver robust evidence to justify their implementation.

The use of a parallel prospective cohort design, and the collection of health data in sync with the implementation of the ULEZ, enables the CHILL study to deliver reliable estimates of exposure-response functions and to inform potential causal relationships between pollutant exposures and children’s health and development. With increasing proportions of the world’s population now living in large urban areas, commonly far exceeding WHO pollution limit guidelines,^3^ CHILL study findings will have important implications for the design and implementation of LEZs and Clean Air Zones in the UK, Europe and beyond.

## Data Availability

Data are available on request to the CI.

## Authors contributions

All authors contributed to writing the protocol. GC, IT, JS, and JC prepared the first draft of the protocol paper, on which all authors commented. IM, FJK and SB received further support from the National Institute for Health Research Health Protection Research Unit (NIHR HPRU) in Environmental Exposures and Health at Imperial College in partnership with Public Health England (PHE). CG and JS receive funding from NIHR ARC North Thames.

## Acknowledgements

We are grateful to all participating schools, parents and children for their enthusiasm and support.

